# EMG-projected MEG High-Resolution Source Imaging of Human Motor Execution: Brain-Muscle Coupling above Movement Frequencies

**DOI:** 10.1101/2023.06.23.23291825

**Authors:** Ming-Xiong Huang, Deborah L. Harrington, Annemarie Angeles-Quinto, Zhengwei Ji, Ashley Robb-Swan, Charles W. Huang, Qian Shen, Hayden Hansen, Jared Baumgartner, Jaqueline Hernandez-Lucas, Sharon Nichols, Joanna Jacobus, Tao Song, Imanuel Lerman, Maksim Bazhenov, Giri P Krishnan, Dewleen G. Baker, Ramesh Rao, Roland R. Lee

## Abstract

Magnetoencephalography (MEG) is a non-invasive functional imaging technique for pre-surgical mapping. However, movement-related MEG functional mapping of primary motor cortex (M1) has been challenging in presurgical patients with brain lesions and sensorimotor dysfunction due to the large numbers of trails needed to obtain adequate signal to noise. Moreover, it is not fully understood how effective the brain communication is with the muscles at frequencies above the movement frequency and its harmonics. We developed a novel Electromyography (EMG)-projected MEG source imaging technique for localizing M1 during ∼l minute recordings of left and right self-paced finger movements (∼1 Hz). High-resolution MEG source images were obtained by projecting M1 activity towards the skin EMG signal without trial averaging. We studied delta (1-4 Hz), theta (4-7 Hz), alpha (8-12 Hz), beta (15-30 Hz), and gamma (30-90 Hz) bands in 13 healthy participants (26 datasets) and two presurgical patients with sensorimotor dysfunction. In healthy participants, EMG-projected MEG accurately localized M1 with high accuracy in delta (100.0%), theta (100.0%), and beta (76.9%) bands, but not alpha (34.6%) and gamma (0.0%) bands. Except for delta, all other frequency bands were above the movement frequency and its harmonics. In both presurgical patients, M1 activity in the affected hemisphere was also accurately localized, despite highly irregular EMG movement patterns in one patient. Altogether, our EMG-projected MEG imaging approach is highly accurate and feasible for M1 mapping in presurgical patients. The results also provide insight into movement related brain-muscle coupling above the movement frequency and its harmonics.

## Introduction

Magnetoencephalography (MEG) is a non-invasive functional imaging technique that directly measures neuronal activity. MEG is useful for presurgical functional mapping of primary motor (M1) cortex due to its excellent temporal (1 millisecond) and spatial resolution (several millimeters) at the cortical level (Huang et al., 2006; Leahy et al., 1998). Critically, MEG recordings of neural activity are not altered by abnormal blood flow in blood-rich tumors or arteriovenous malformations, unlike the blood-oxygen-level-dependent (BOLD) signal in fMRI-based functional mapping of pre-surgical cases. Still, movement-related functional mapping using MEG has been challenging in patients with brain lesions or stroke with sensorimotor dysfunction because large numbers of trials are needed to generate averaged responses with an adequate signal to noise ratio (SNR). To generate good SNR from MEG-related movement responses, the patient needs to lift his/her finger quickly, with a large displacement, for hundreds of trials in a time-locked fashion. Consequently, the acquisition time for conventional movement paradigms is usually 10-20 minutes (Bowyer et al., 2020; Bulubas et al., 2020; Cheyne et al., 2008, 2006; Gaetz et al., 2010; Huang et al., 2004; Spooner et al., 2022; Tarapore et al., 2012). However, in clinical medical practice, many patients with sensorimotor dysfunction cannot perform several hundred movement repetitions because their finger movements are slow, weak, and not time-locked to movement-related M1 brain activity. Hence, there is an urgent need for quick, accurate, and reliable MEG techniques for movement-related functional mapping within single trials.

The development of a fast, accurate, and reliable MEG-presurgical mapping approach has been partly hindered by an insufficient understanding of the frequency characteristics of movement-related M1-muscle communication. Different modeling approaches and frequency bands have been employed to map M1 cortex activity. For example, MEG event-related desynchronization (ERD) localized M1 cortex activity in the beta band (15-30 Hz) using a beamformer-based spatial filter (Bulubas et al., 2020; Tarapore et al., 2012) whereas MEG evoked-related field (ERF) components localized M1 cortex activity within a broad frequency band setting (e.g., DC to 30 Hz or 100 Hz) using dipole modeling (Bowyer et al., 2020; de Tommaso et al., 2020; Huang et al., 2004; Spooner et al., 2022) and beamformer (Cheyne et al., 2006; Gaetz et al., 2010) approaches. MEG gamma-band M1 activity was mainly found post-movement-onset using a seed virtual sensor placed at M1, which was first located from evoked-related finger movement components (Cheyne et al., 2008; Huo et al., 2010). Yet it is unclear if gamma-band activity can be used *directly* to accurately localize M1 cortex activity. Regardless of the modeling approach, however, few studies have systematically examined movement-related M1 cortex activity across different frequency bands, including lower bands, which influence M1 cortex activity (e.g., Popovych et al, 2016). Moreover, M1 cortex activity typically is not just limited to muscle activity, and non-motor brain activities from background and artifacts may affect brain-muscle communication (Bu et al., 2023).

In contrast, during sustained / steady-state or isometric muscle contraction (SMC), many studies have examined corticomuscular coupling (CMC) between M1 cortex and muscle activity from electromyography (EMG) recordings (see review in (Bourguignon et al., 2019)). We refer to this as SMC-CMC. For example, EMG-MEG coherence while squeezing a force transducer between the fingers is mainly in the beta band (15–30 Hz) and is predominantly driven by the M1 cortex (Grosse et al., 2002; Pohja et al., 2005). Electroencephalography (EEG) studies also report EMG-EEG coherence in the beta band using various SMC tasks (Bayraktaroglu et al., 2011; Ushiyama et al., 2017; Yao et al., 2007; Zheng et al., 2016). However, movement-related functional mapping is the paradigm most commonly employed in clinical practice.

Movement-related CMC (MR-CMC) is not well understood. Rather, most studies focus on cortico-kinematic coupling (CKC) between primary sensorimotor (SM1) cortex and kinematic variables such as the speed, velocity, acceleration, and force of movements (Bourguignon et al., 2019; Jerbi et al., 2007; Piitulainen et al., 2013). CKC predominates at the SM1 cortex and occurs at the movement frequency and its harmonics (see review in (Bourguignon et al., 2019)). Consequently, CKC is thought to be mainly driven by movement rhythmicity during active, passive, and observed movements rather than coherence with muscle activity per se (Bourguignon et al., 2019). When mapping M1 function during voluntary movements, movement-related brain-EMG coupling and MR-CMC in lower frequency bands such as delta (1-4 Hz) and theta (4-7 Hz) are not completely understood. In particular, it is unclear if strong brain-EMG coupling exists in MEG *frequency bands that are above the movement frequency and its harmonics*. This knowledge gap is puzzling since studies of patients with tremors found significant EMG-EEG and EMG-MEG coherence from M1 cortex, which peaked in low-frequency bands (e.g., delta and theta) (Hallett et al., 2021; Timmermann et al., 2003).

To address these limitations, the present study developed a novel approach based on EMG-projected MEG high resolution source imaging of self-paced, repetitive index finger movements. In this approach, cortical MEG signals were directly projected to the parameter space expanded by the EMG activity, which substantially reduced the effects of non-movement related brain activity and artifacts. Unlike conventional MEG movement tasks, which require many repetitions and take a long time to perform, our task lasted about 1 minute and required participants to perform continuous self-paced, index finger flexions and extensions at ∼ 1 Hz repetition rate, which can usually be performed by brain tumor or stroke patients with sensorimotor weakness. Our first aim was to evaluate the accuracy of this new EMG-projected MEG source imaging analysis for localizing M1 cortex in healthy adults. In this regard, we sought to identify M1 cortex frequency bands (delta, theta, alpha, beta, gamma) that communicate with the muscle in individual subjects. The theta band was of particular interest, since in our study potential brain-EMG coupling in the theta band is above the movement frequency (∼1 Hz) and its harmonics. A second exploratory aim was to test the feasibility and efficacy of this approach in localizing contralateral M1 cortex activity in two clinical patients with sensorimotor weakness due to tumor or brain injury.

## Method and Materials

### Participants

The EMG-projected MEG approach was first evaluated by testing thirteen right-handed healthy participants who were free of neurological disorders (10 males, 3 females, age range of 19-51, mean 32.9 ± 10.4). Next, we sought to evaluate the precision of this approach for localizing brain areas that were activated by finger movements in two patients with brain lesions near the motor and somatosensory areas. Patient 1 was a right-handed male in the age range of 50-55 with history of metastatic melanoma who presented with a right facial droop and problems with right-sided coordination and fine-motor movement. MRI revealed a left posterior-frontal intrinsically T1-bright mass in the left precentral gyrus in vicinity of hand locus, and another similar mass in the right middle temporal gyrus, consistent with the subsequent surgically-proven diagnosis of metastatic melanoma. Patient 2 was a left-handed male in the age range of 15-20 with a history of perinatal stroke, right hemiplegic cerebral palsy, and symptomatic intractable focal epilepsy. He has chronic right upper extremity numbness with mild weakness around the right forearm/hand. An MRI exam showed chronic cystic encephalomalacia from perinatal stroke, involving the anterior-inferior parietal lobes, with the left much worse than right, primarily involving the lateral left peri-rolandic region and the adjacent more posterior left parietal lobe. Secondary thinning of the corpus callosum was also present.

The study protocols were approved by institutional review boards of the VA San Diego Healthcare System and University of California, San Diego. The healthy participants gave written informed consent prior to study procedures. The informed consent followed the ethical guidelines of the Declarations of Helsinki (sixth revision, 2008). The clinical patients signed the HIPPA waiver form and/or COTA which allowed their data to be used for research and educational purposes.

### Self-Paced Finger Movement Task and EMG Recordings

During MEG recordings, the participant was seated with his/her left and right arms resting on a table that was positioned in front of the body; the palm was facing up when the subject performed index finger movements. Two pairs of bipolar surface EMG electrodes were placed on the forearm to record the EMG signals. The reference electrodes from both pairs were placed at the wrist (touching each other), whereas the non-reference electrodes were placed on the extensor digitorum and flexor digitorum superficialis (Andrews et al., 2009). Subjects performed a finger movement task in which they were instructed to flex and extend their index finger (**Fig. 1, top panel insert**) continuously in a self-paced manner at ∼ 1 Hz rate for about 60 seconds. Left and right index finger movements were performed separately, and their order was counterbalanced across participants. Movement-related EMG signals were recorded simultaneously with the MEG signals.

**Figure 1:**
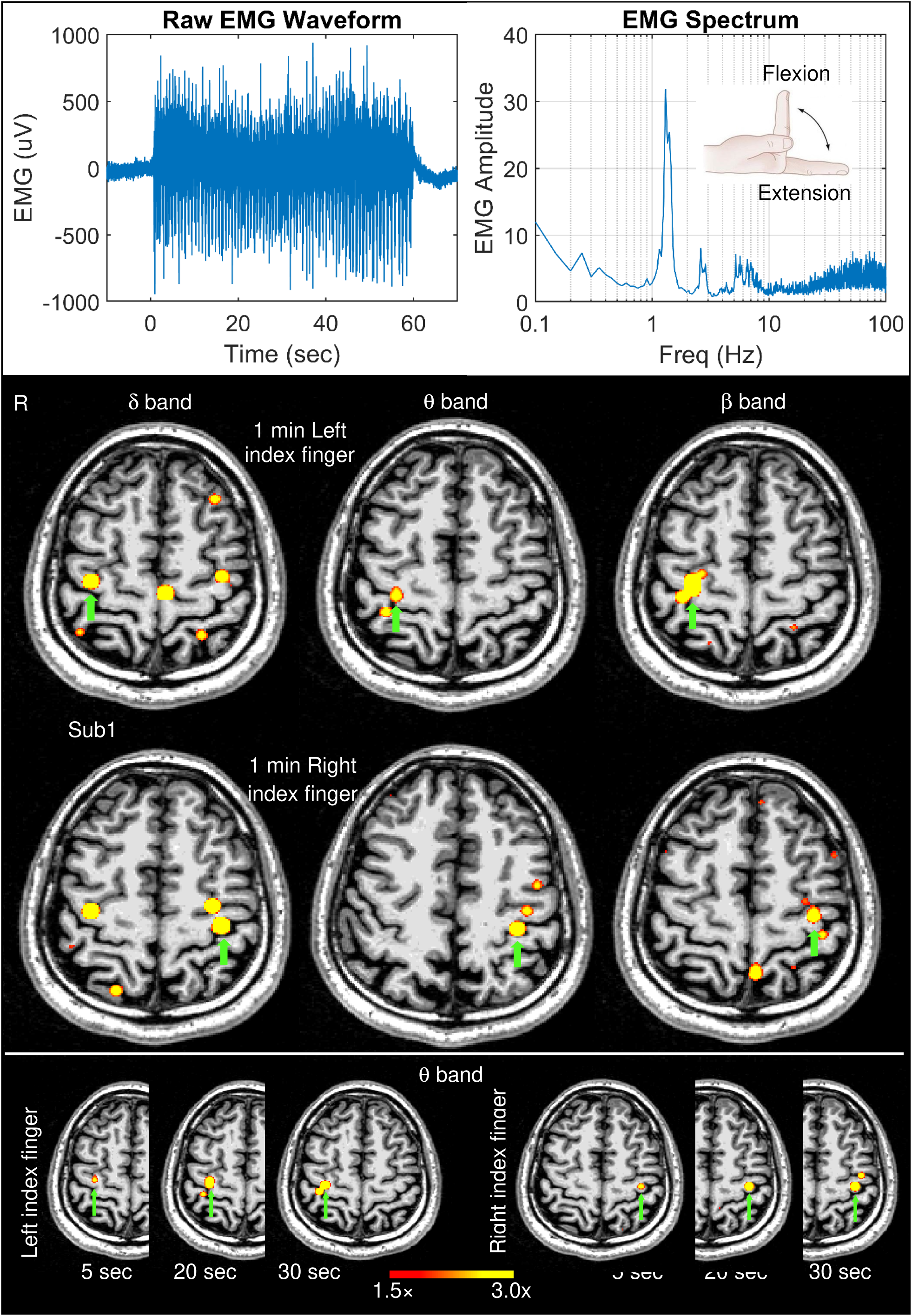
Movement-related EMG signals and EMG-projected MEG source images for a healthy subject. Top panel: The left graph displays the EMG waveform of a surface electrode during the self-paced (∼1 min) index finger movement in Subject 1 (Sub 1). The right graph displays the EMG spectrum which shows the amplitude of a muscle activity as a function of frequency. The insert illustrates an index finger flexion and extension during self-paced movements. Middle panel: Significant primary motor cortex sources (green arrows) contralateral to left finger and right self-paced index finger movements for delta, theta, and beta bands in Sub 1. Bottom panel: Theta band source images for the first 5 sec, 20 sec, and 30 sec time windows during left and right finger movements. The color bar shows the activity threshold at ≥ 1.5× of the empty room maximum value, and a saturation level at 3.0× for images in the middle and bottom panels.

### MEG Data Acquisition and Preprocessing

MEG motor responses (spontaneous recording) and the associated movement-related EMG signals were collected at the UCSD MEG Center using a 306-channel whole-head VectorView™ MEG system (MEGIN-Neuromag, Helsinki, Finland). Participants sat inside a multi-layer magnetically-shielded room (IMEDCO-AG) (Cohen et al., 2002). Precautions were taken to ensure head stability; foam wedges were inserted between the participant’s head and the inside of the unit, and a Velcro strap was placed under the participant’s chin and anchored in superior and posterior axes. Head movement across different sessions was about 2-3 mm. Data were sampled at 1000 Hz and were run through a high-pass filter with a 0.1 Hz cut-off, and a low-pass filter with a 300 Hz cut-off. The filter associated with MEG data acquisition is a first-order time-domain filter with 3 dB around the cut-off points. Eye blinks, eye movements, and heart signals were monitored. Sixty seconds of empty room data were also collected to control for background noise.

MEG data were first run through MaxFilter, also known as signal space separation (Song et al., 2009; Taulu et al., 2004; Taulu and Simola, 2006) to remove external sources of interference (e.g., magnetic artifacts due to metal objects, strong cardiac signals, environment noises, etc.). Next, residual artifacts due to eye movements, residual cardiac signals, and 60 Hz powerline artifacts were removed using Independent Component Analysis using Fast-ICA (Hyvärinen, 1999; Hyvärinen and Oja, 2000).

### MRI

A structural MRI of each healthy participant’s head was collected on a General Electric 1.5T Excite MRI scanner using a standard high-resolution anatomical volume with a resolution of 0.94×0.94×1.2 mm3 and a T1-weighted 3D-IR-FSPGR pulse sequence.

The two clinical patients had standard clinical T1- and T2-weighted, FLAIR, diffusion-weighted, and susceptibility-weighted sequences, as well as postcontrast T1-weighted images. In addition, Patient 1 had postcontrast axial 3D 1.1-mm T1-weighted VIBE, and Patient 2 had axial non-contrast 3D 1.2-mm T1-weighted BRAVO, which were used for co-registration with the MEG data.

### MEG-MRI Registration and BEM Forward Calculation

To co-register the MEG with MRI coordinate systems, three anatomical landmarks (i.e., left and right pre-auricular points, and nasion) were measured for each participant using the Probe Position Identification system (Polhemus, USA). By using MRILAB (MEGIN/Neuromag) to identify the same three points on the participant’s MR images, a transformation matrix involving both rotation and translation between the MEG and MR coordinate systems was generated. To increase the reliability of the MEG-MR co-registration, approximately 100+ points on the scalp were digitized with the Polhemus system along with the three landmarks; these points were co-registered onto the scalp surface of the MR images.

The T1-weighted images were also used to extract the brain volume and innermost skull surface (SEGLAB software developed by MEGIN/Neuromag). Realistic Boundary Element Method (BEM) head model was used for MEG forward calculation (Huang et al., 2007; Mosher et al., 1999). The BEM mesh was constructed by tessellating the inner skull surface from the T1-weighted MRI into ∼6000 triangular elements with ∼5 mm size. A cubic source grid with 5 mm size was used for calculating the MEG gain (i.e., lead-field) matrix, which leads to a grid with ∼10,000 nodes covering the whole brain.

### EMG-Projected MEG Source Imaging Solution

The novel EMG-projected MEG source modeling developed in the present study was based on an enhanced version of the Fast-VESTAL algorithm and included EMG signal projection. Fast-VESTAL and VESTAL algorithms published previously (Huang et al., 2016, 2014a, 2006) provide high-resolution MEG source images for resting-state and evoked paradigms (Edgar et al., 2022; Huang et al., 2016, 2020b, 2020a, 2019, 2017, 2014a, 2014b, 2014c; Ji et al., 2022). In the present study, an enhanced version of Fast-VESTAL formulations by the primary developer (M.X. Huang) adopted a generalized second order cone programming (GSOCP) for the L1 minimum norm solver. The enhanced Fast-VESTAL has been independently validated by other laboratories who reported good performances against other state-of-the-art MEG source imaging techniques (e.g., (Zheng et al., 2021)). The new theoretical formulation of the high-resolution EMG-projected MEG source imaging approach is presented here.

#### System Equation

First, we take an imaging (lead-field) approach and divide the source space (gray-matter brain volume) into a grid with several thousand nodes. An electrical current dipole is assigned to each node. MEG time-domain sensor-waveform signals can then be expressed in a data matrix: **B**=[**b**(*t_1_*),**b**(*t_2_*),…, **b**(*t_T_*)], where *t_1_, t_2_, …, t_T_* are time samples and *T* is the total number of time samples and **b**(*t_i_*) is a *M×1* vector containing the magnetic fields at *M* sensor sites at time sample *ti*. This *M×T* data matrix can be expressed as the system equation:

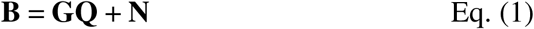

where **G** is an *M×2P* gain (lead-field) matrix calculated from MEG forward modeling for the pre-defined source grid with *P* dipole locations, with each dipole location having two orthogonal orientations (i.e., θ and φ). **N** is an *M×T* noise matrix. **Q** is a *2P×T* source time-course matrix. In the spherical MEG forward head model, θ and φ represent the two tangential orientations for each dipole location, whereas in a realistic MEG forward model using the BEM, the θ and φ-orientations are obtained as the two dominant orientations from the singular-value decomposition (SVD) of the *M×3* lead-field matrix for each dipole, as previously documented (Huang et al., 2006). The noise term in Eq. (1) is assumed to be Gaussian white noise. If correlated noise exists, an automated pre-whitening procedure can be applied (Huang et al., 2014a). The inverse solution in Eq. (A1) obtains the source time-courses **Q** for given MEG sensor waveforms **B**.

#### EMG-projected MEG signals with time delays

Now, introduce a matrix that contains the EMG signal matrix from an EMG electrode with *D* different time delays:

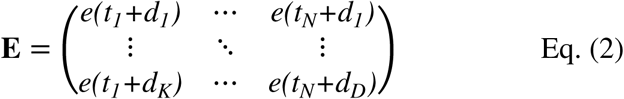

where *e(t_i_+d_j_)* are normalized EMG signals with time delay *d_j_*, in relation to the MEG time samples *t_i_*. In the present study, we are interested in -100 ms to 0 ms time window, which marks the beginning of the motor execution phase. By projecting MEG signal **B** toward the EMG signal **E**, we obtain:

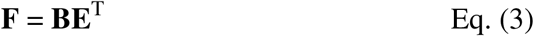

where F is the EMG-projected MEG sensor signals with dimensions *M×D.* Essentially, Eq. (3) is the EMG-projected MEG sensor waveform for different time delays. The following formulation is to find the Fast-VESTAL inverse source imaging solution for Eq. (3).

In the Fast-VESTAL approach, we first remove the time-delay-dependent features from Eq. (3) and only focus on the spatial profiles. This is done by performing a SVD for the *M×D* EMG-projected MEG sensor waveform data matrix:

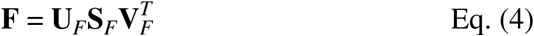

The dimensions for **U***_F_*, **S***_F_*, and **V***_F_*are *M×M, M×D,* and *D×D,* respectively. All time-delay information in the MEG sensor waveform can be represented as a linear combination of the singular vectors in the matrix **V***_F_*. In addition, SVD is performed for the gain matrix **G**:

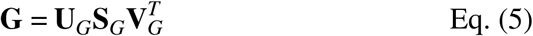

The dimensions for **U***_G_*, **S***_G_*, and **V***_G_*, are *M×M, M×2P,* and *2P×2P,* respectively. Substituting Eqs. (4)(5) into Eq. (3) and then performing an operation by multiplying the result with **V***_F_* from the right side, we have:

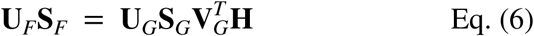

The *2P×M* matrix **H** = **QV***_F_* is called the EMG-projected MEG <u>source delay map</u> matrix for the given time-delay window and is *independent* of individual time-delay samples. In the above derivation, we also make use of the fact that the white noise is uncorrelated with the MEG neuronal signals **NV***_F_* **= 0**.

Each column of **U***_F_***S***_F_* is defined as a <u>spatial mode</u> of MEG sensor-waveforms. The significance of Eq. (6) is that each spatial mode in the sensor-waveforms be expressed as a linear combination of the corresponding source imaging maps (i.e., the columns of **H**). It is clear that the number of signal (i.e., dominant) spatial modes in a given MEG data set (usually ranges from 1--10) is substantially less than the number of time-delay samples in the data (∼200). Thus, by solving Eq. (6), the computational cost can be substantially reduced.

#### Fast-VESTAL Minimum L1-norm Solution using GSOCP

Eq. (A4) is under-determined, with the number of unknown variables in each column of **H** = [**h***_1_*,**h***_2_*,…,**h***_k_*,…,**h***_M_*] (i.e., *2P*) much larger than the number of sensor measurements in each column of **U***_F_***S***_F_*= [*s_1_***u***_1_*, *s_2_***u***_2_*,…,*s_k_***u***_k_*,…,*s_M_***u***_M_*] (i.e., *M*), so additional constraint(s) are needed to obtain a unique solution for Eq. (6). Furthermore, the number of signal (dominant) spatial modes *k* is usually much smaller than the number of MEG sensor measurements *M*. After multiplying from the left side with 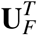, for individual dominant spatial modes of Eq. (6), Eq. (6) can be written as:

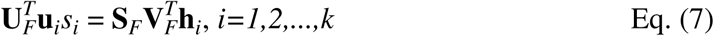

where *i=1,2,…,k* are the indices of spatial modes in sensor space, and the *2P×1*vector **h***_i_* is the source imaging map associated with the dominant spatial mode vector **u***_i_* (dimension *M×1*) of the sensor-domain waveforms.

Eq. (7) is still underdetermined and an additional constraint is needed in order to obtain a unique solution. In the present study, GSOCP is used to solve Eq. (7). In this approach, the Fast-VESTAL minimum L1-norm solution **h***_i_* of Eq. (7) is:

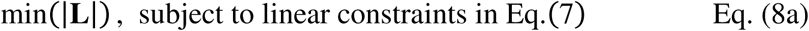

where **|L|** is the L1-norm with GSOCP:

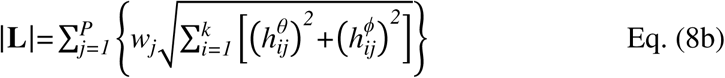

In Eq. (8b), 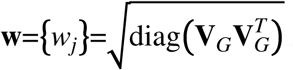 is a *2P×1* weighting vector was chosen to optimally remove bias towards grid nodes at the superficial layer and obtain accurate localization in depth (U.S. Patent, Provisional Application Attorney Docket No.: 009062-8264.US00) (Huang et al., 2014a). In conventional minimum L1-norm solutions, there is a bias associated with source orientations. In general, the solution is in favor of activity along the principal axes (i.e., *θ*^^^ and *φ*^^^) of the dipole moments at the *j^th^* source node: 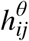 and 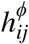. Here, such bias is directly removed which makes minimum L1-norm solution rotational invariance using the second-order cone programming (SOCP), similar to others (Haufe et al., 2011; Huang et al., 2016; Ou et al., 2008). And here, SCOP was generalized across *k* dominant spatial modes in sensor space using the 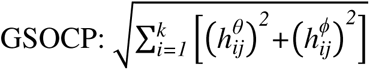. SeDuMi software (http://sedumi.ie.lehigh.edu/) was used as the L1-norm solver.

After solving for **h***_i_* and hence **H**, the voxel-wise Fast-VESTAL source imaging result can be obtained on the source grid as the source magnitude vector:

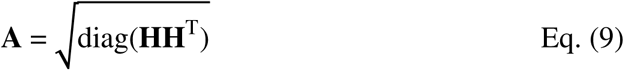

which is the *2P×1* source magnitude value across grid nodes. The main feature of source magnitude vector **A**, the EMG-projected MEG source imaging solution using Fast-VESTAL with GSCOP, is that it is highly sparse, with many of its elements being either zero or close to zero, as a direct consequence of L1-norm minimization. An objective pre-whitening method was applied to remove correlated sensor and environmental noise and objectively select the dominant eigen-modes (i.e., *k*) of the sensor-waveform covariance matrix (Huang et al., 2014a).

If more than one EMG channel is used, the above procedure (i.e., Eqs. (1) - (9)) will be repeated for each EMG channel separately. Then at the end, the source magnitude vectors are combined in the form of the Euclidean norm to assemble the final voxel-wise Fast-VESTAL source imaging map.

### EMG-Projected MEG Source Imaging: Finger Movement Task

We first analyzed the EMG-projected MEG signals from ∼60-second left or right index finger movements in the 13 healthy subjects. To systematically study the nature of brain-muscle communication across different frequency bands, EMG-projected MEG source images were examined for delta (1-4 Hz), theta (4-7 Hz), alpha (8-12 Hz), beta (15-30 Hz), and gamma (30-90 Hz) frequency bands. In each subject, the voxel-wise MEG magnitudes (Eq. 9) related to the two EMG surface electrode channels were combined using the Euclidean norm. The time delay variable was chosen to be from -100 ms to 0 ms, which marks the beginning of the early motor execution phase. The voxel-wise maps with the maximum magnitude across different delays for each voxel were shown in the figures of the Results section.

The same approach was applied to empty-room data, which also lasted ∼ 60 seconds. M1 cortex activity was assessed using a conservative thresholding approach wherein the maximum MEG source magnitude from the empty-room data for all voxels at the cortical level was obtained. Then, a threshold was chosen as ≥ 1.5× of the maximum MEG source magnitude value from the empty-room data. This threshold was used to display EMG-projected MEG source images during the self-paced finger movement task for both healthy controls and patients.

## Results

### Movement-related EMG waveform and spectrum

**Fig. 1 (top panel, left graph)** shows the EMG waveform of a surface electrode from a representative healthy subject (Subject 1) during the ∼1 min self-paced index finger movement. The right graph shows the spectrum of the EMG data obtained from the Fast Fourier Transform (FFT) by averaging the absolute value of the FFT signals across moving time windows, each with 20-sec duration and 1-ms time increments across the ∼1 min duration. The fundamental frequency at 1.30 Hz is the dominant peak in the spectrum and its much weaker second harmonics at 2.60 Hz is also visible. There was a broader peak ∼4-8 Hz which is above the movement frequency and its harmonics, followed by some high frequency components. Across all 13 healthy subjects, the frequencies of the main peaks in the EMG spectra during the left and right finger movements were 1.33 ± 0.44 Hz and 1.29 ± 0.35 Hz, respectively, which did not differ significantly (paired t-test: p = 0.44).

### Movement-related EMG-projected MEG source images

**Fig. 1** (middle panel) shows the EMG-projected MEG source images from left and right self-paced (∼1 min) index finger movements for delta, theta, and beta bands. In all cases, M1 cortex activity contralateral to the side of the finger movement (green arrows) was accurately localized. For the delta band, but not theta or beta bands, M1 cortex activity ipsilateral to the finger movement also showed significant activity. The result from Subject 1 clearly shows that EMG-projected MEG source imaging accurately localizes M1 cortex delta, theta, and beta band activities. Contralateral M1 cortex activity for alpha and gamma bands was below threshold.

To explore the limit of this approach, finger-movement time windows as short as 30-sec, 20-sec, and even 5-sec from the beginning of the movement were used for the creating the EMG-projected MEG source images for the theta band. Here, we focused on theta band since 4-7 Hz is completely outside the base frequency of the ∼1 Hz finger movement and its harmonics, rendering it unlikely that the base movement frequency would contaminate theta band activity. **Fig. 1** (bottom panel) shows that M1 cortex activity contralateral to the finger movement was accurately obtained in each of these time windows, even the 5-sec window. The source images for the 20- and 30-sec time windows were highly similar to the results from the 1-min time window (middle panel). The volume of significant activation for the 5-sec time window was lower, resulting from lower SNR due to fewer movement repetitions in a very short time duration.

### Group results for delta band

**Fig. 2** shows the EMG-projected MEG source activity in the *delta* band from the remaining 12 healthy subjects for the self-paced finger movements (∼1 min). For both left and right index finger movements, M1 cortex activity contralateral to movement was significant in all 24 cases (green arrows) as it was for Subject 1 (**Fig. 1**). Thus, in 100% of the 13 healthy subjects (26 cases), contralateral M1 delta-band activity was obtained for EMG-projected MEG source images. Ipsilateral M1 cortex also showed delta activities in Subject 3 (right finger movements) and Subject 8 (left finger movement), similar to Subject 1 (**Fig. 1**, middle panel).

**Figure 2:**
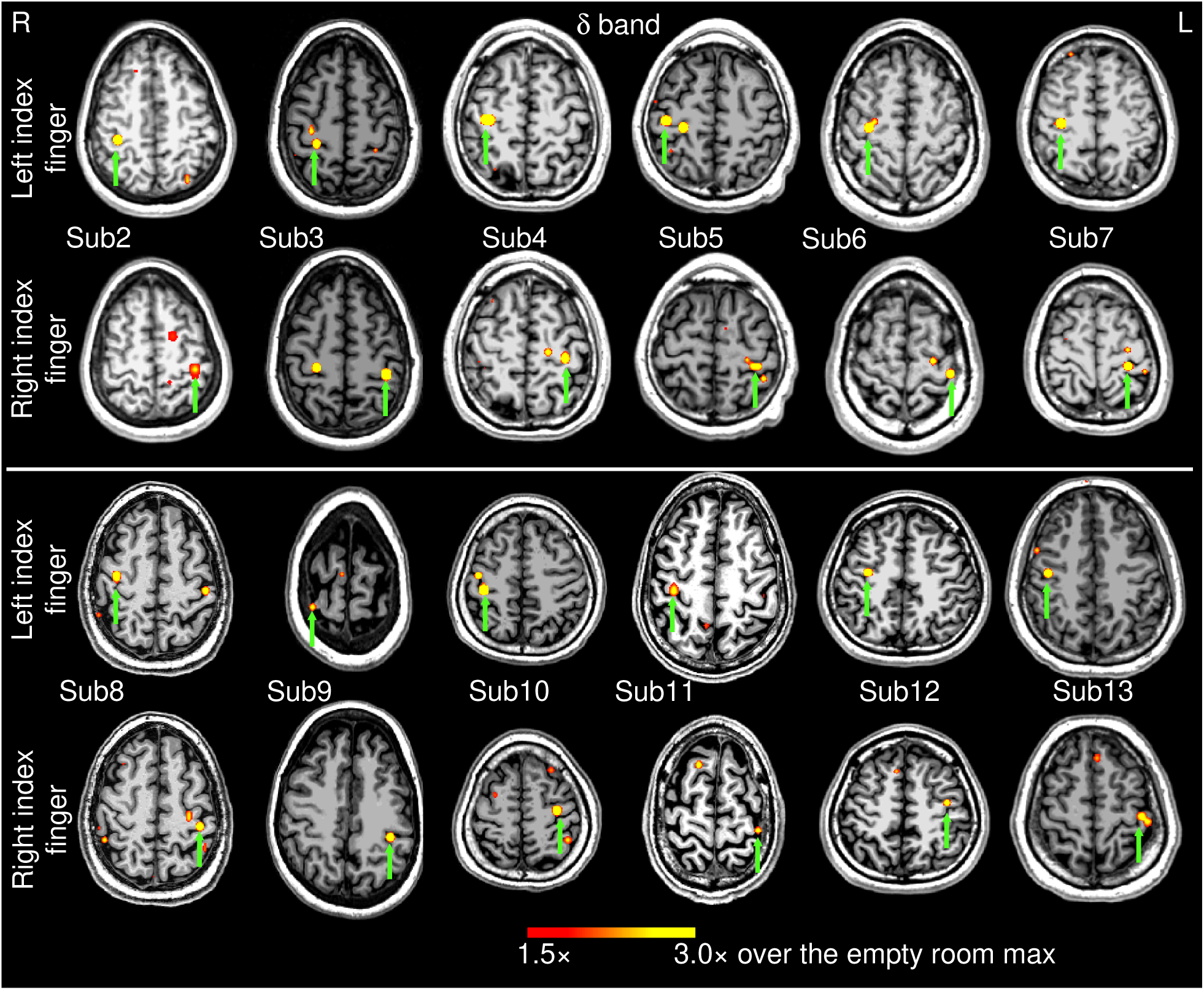
Movement-related delta-band EMG-projected MEG source images for the remaining 12 healthy subjects. Significant primary motor cortex sources (green arrows) contralateral to the left or right self-paced (∼1 min) index finger movements. The color bar shows the activity threshold at ≥ 1.5× of the empty room maximum value, and a saturation level at 3.0×. Sub = subject

### Group results for theta band

**Fig. 3** shows the EMG-projected MEG source images in the *theta* band from the 12 healthy subjects during self-paced finger movements (∼1 min). Like the delta-band results, M1 cortex activity contralateral to both left and right index finger movements was significant in all 24 cases (green arrows), as it was for Subject 1 (**Fig. 1**, middle panel).

**Figure 3:**
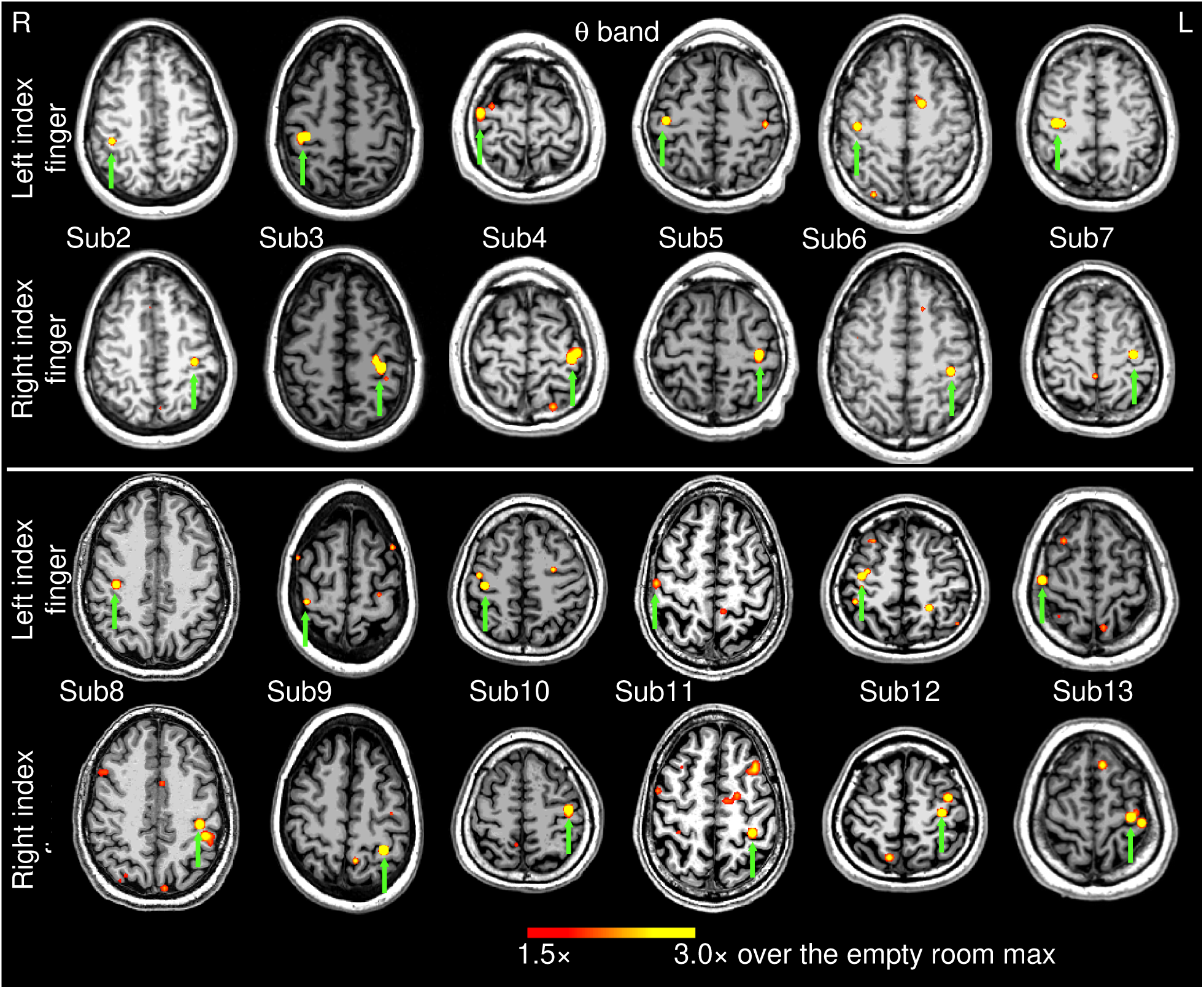
Movement-related theta-band EMG-projected MEG source images for the remaining 12 healthy subjects. Significant primary motor cortex sources (green arrows) contralateral to the left or right self-paced (∼1 min) index finger movements. The color bar shows the activity threshold at ≥ 1.5× of the empty room maximum value, and a saturation level at 3.0×. Sub = subject

For exploratory purposes, we also analyzed the data using the MEG response from EMG activity for the first 30-sec and 5-sec time windows of self-paced finger movements in all 13 subjects. For the 30-sec time window, the EMG-projected MEG source imaging approach accurately localized the contralateral M1 cortex in 25 out of 26 (or 96.2%) cases. The source locations were virtually the same as in **Fig. 3** and **Fig. 1** (bottom panel), hence we did not show their source locations. Even for the 5 sec duration, EMG-projected MEG source imaging approach localized the contralateral M1 cortex in 19 out of 26 (or 73.1%) cases. However, more spontaneous brain activities seemed to be present which may contaminate the results (see Discussion). Thus, going forward, the remaining analyses of healthy subjects focused on the 1-min movement duration.

### Group results for beta band

**Fig. 4** shows the EMG-projected MEG source images in the *beta* band from the 12 healthy subjects self-paced finger movements (∼1 min). In 18 out of the 24 (74%) left and right index finger cases, contralateral M1 cortex activity (green arrows) was significant. However, in 6 cases, contralateral MEG activities did not reach threshold for either left or right finger movement (Subjects 2 and 8), or just for the right finger movement (Subjects 12 and 13). Including Subject 1 from **Fig. 1**, contralateral M1 activity was obtained in 20 out of 26 cases (or 76.9%) with beta-band EMG-projected MEG source imaging.

**Figure 4:**
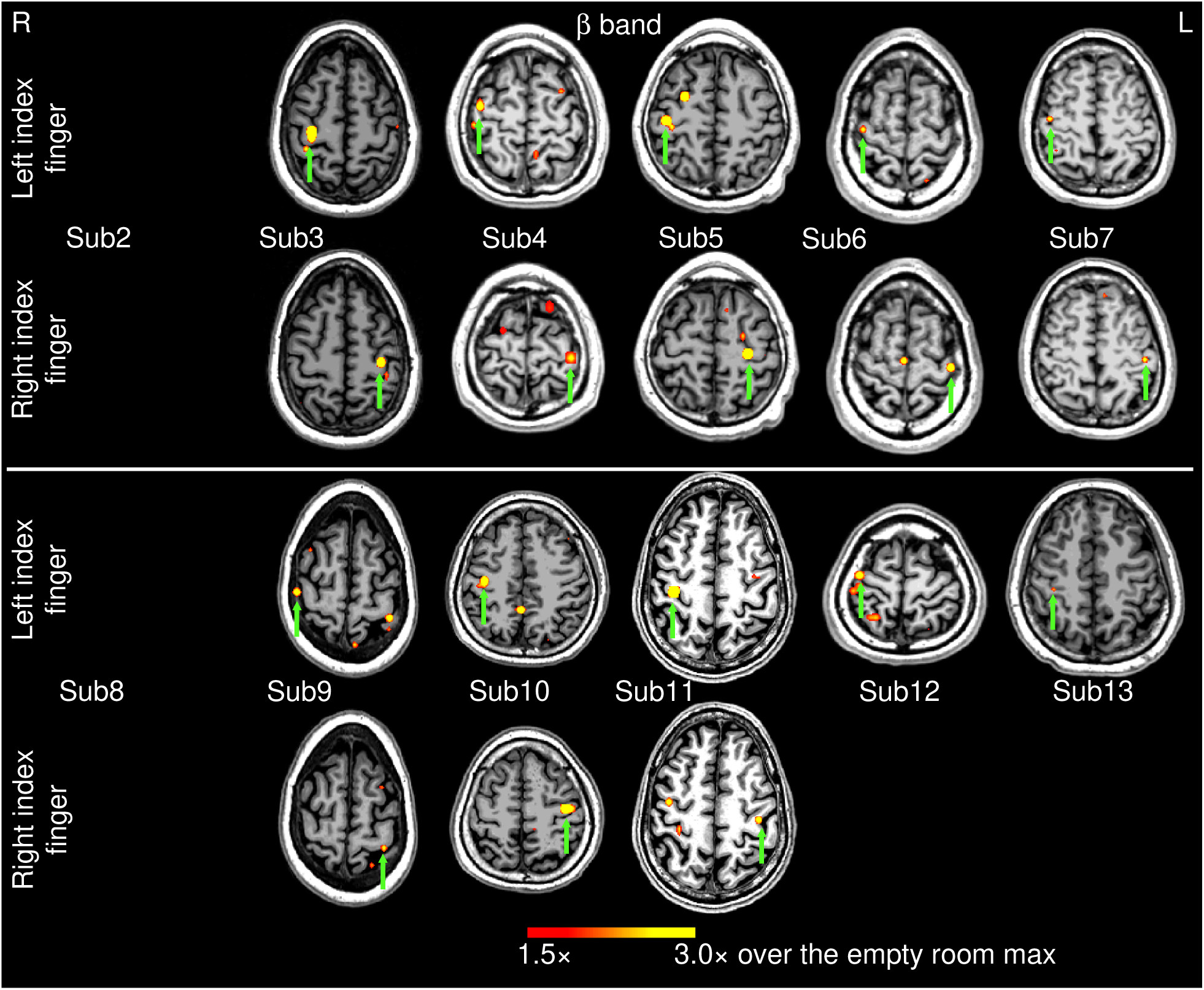
Movement-related beta-band EMG-projected MEG source images for the remaining 12 healthy subjects. Primary motor cortex sources (green arrows) contralateral to self-paced (∼1 min) left or right index finger movements were significant in 18 out of 24 cases. The color bar shows the activity threshold at ≥ 1.5× of the empty room maximum value, and a saturation level at 3.0×. In 6 cases, contralateral primary motor source activity did not reach threshold. Sub = subject

### Group results for alpha and gamma bands

**Fig. S1** in Supplementary Material shows the EMG-projected MEG source images in the *alpha* band from the 12 healthy subjects for self-paced (∼1 min) finger movements. In only 9 out of the 24 left and right finger movement cases, M1 cortex activities contralateral to the finger movement were significant (green arrows). As contralateral MEG activities in Subject 1 (left and right movements) also did not pass the threshold, contralateral beta-band M1 activity was obtained in 9 out of 26 cases (or 34.6%) using EMG-projected MEG source imaging. Unlike the other frequency bands, contralateral M1 cortex activity for *gamma*-band signals was nonsignificant in all 26 finger movement cases.

### Coordinates of M1 sources in the standard MNI-152 space

We also co-registered the EMG-projected MEG source images to the MNI-152 (Grabner et al., 2006) brain atlas template using a linear affine transformation program, FLIRT, from FSL software (www.fmrib.ox.ac.uk/fsl/) (Smith et al., 2004; Woolrich et al., 2009). The source coordinates for alpha band were not included due to large percentage of cases below the thresholds. **Table 1** shows the overall M1 source locations across the delta, theta, and beta frequency bands are similar for the left and right index finger movements. Only the x coordinates for the right contralateral M1 showed larger values in theta than the delta band (t = 2.6, p < 0.05, df = 24, uncorrected for multiple comparisons), and no other coordinates showed statistical differences.

**Table 1:** MNI-152 coordinates (x, y, and z in mm) of primary motor cortex contralateral to left or right finger movements, for delta, theta, and beta bands.

| Subject ID | L-Finger, R-M1, δ-band | R-Finger, L-M1, δ-band | L-Finger, R-M1, θ-band | R-Finger, L-M1, θ-band | L-Finger, R-M1, β-band | R-Finger, L-M1, β-band |
| --- | --- | --- | --- | --- | --- | --- |
| 1 | 37, -10, 57 | -34, -21, 53 | 32, -20, 57 | -33, -20, 52 | 32, -10, 57 | -33, -10, 57 |
| 2 | 34, -20, 47 | -38, -25, 57 | 42, -25, 57 | -38, -20, 47 | --- | --- |
| 3 | 27, -25, 57 | -33, -30, 52 | 37, -20, 52 | -33, -20, 52 | 32, -15, 57 | -33, -15, 57 |
| 4 | 37, -25, 57 | -33, -30, 57 | 42, -20, 67 | -38, -30, 62 | 37, -15, 58 | -38, -30, 67 |
| 5 | 42, -25, 57 | -33, -35, 57 | 42, -25, 57 | -38, -25, 57 | 42, -25, 57 | -28, -25, 57 |
| 6 | 32, -15, 57 | -38, -25, 67 | 42, -15, 52 | -38, -25, 52 | 37, -15, 67 | -38, -20, 67 |
| 7 | 32, -20, 52 | -28, -30, 62 | 37, -20, 52 | -33, -20, 57 | 42, -15, 57 | -43, -25, 52 |
| 8 | 37, -5, 52 | -38, -20, 57 | 37, -15, 42 | -38, -20, 47 | --- | --- |
| 9 | 32, -20, 72 | -38, -20, 47 | 37, -20, 67 | -33, -30, 57 | 47, -10, 62 | -33, -25, 67 |
| 10 | 37, -25, 57 | -33, -15, 58 | 37, -20, 57 | -39, -19, 56 | 42, -20, 57 | -37, -21, 55 |
| 11 | 37, -15, 52 | -38, -15, 67 | 46, -6, 57 | -33, -20, 57 | 37, -15, 52 | -38, -10, 52 |
| 12 | 32, -10, 47 | -38, -10, 52 | 37, -15, 57 | -33, -25, 57 | 37, -10, 67 | --- |
| 13 | 42, -15, 47 | -43, -15, 57 | 47, -10, 62 | -33, -15, 62 | 37, -20, 52 | --- |
| mean | 35.2, -17.7, 54.7 | -35.8, -22.4, 57.2 | 39.6, -17.8, 56.6 | -35.4, -22.2, 55.0 | 38.4, -15.5, 58.5 | -35.7, -20.1, 59.0 |
| SD | 4.3, 6.7, 6.7 | 3.8, 7.5, 5.7 | 4.3, 5.4, 6.6 | 2.7, 4.4, 4.8 | 4.5, 4.7, 5.0 | 4.4, 7.1, 6.3 |

### EMG-projected MEG source imaging for clinical patients

Since M1 source localization in the theta band (4-7 Hz) was highly accurate (100%) and fell outside of the ∼1 Hz fundamental frequency of self-paced finger movements, EMG-projected MEG source imaging was applied to the theta band in two patients with brain lesions near the central sulcus using the same threshold setting as for the healthy subjects. Patient 1 had a left posterior frontal metastatic melanoma. **Fig. 5** (top panel, left plots) shows the EMG waveform and its spectrum during right index finger self-paced movements (∼ 1 min). The spectrum’s main peak at 1.35 Hz was the fundamental movement frequency, followed by a weaker second harmonic around 2.7 Hz and higher frequency activities. The middle panel of **Fig. 5** shows that his contralateral left hemisphere M1 source activity was accurately localized anterior to the distorted left central sulcus, just posterior to the mass in the left precentral gyrus. As expected, M1 activity in the non-lesioned right hemisphere was significant, and the EMG waveform and spectrum were like those of healthy subjects. (**Supplementary Fig. S2**, middle and bottom panels). In both hemispheres, theta band M1 source activity was also accurately localized for the 30 sec and 5 sec time windows (**Supplementary Fig. S2**, top and bottom panels).

**Figure 5:**
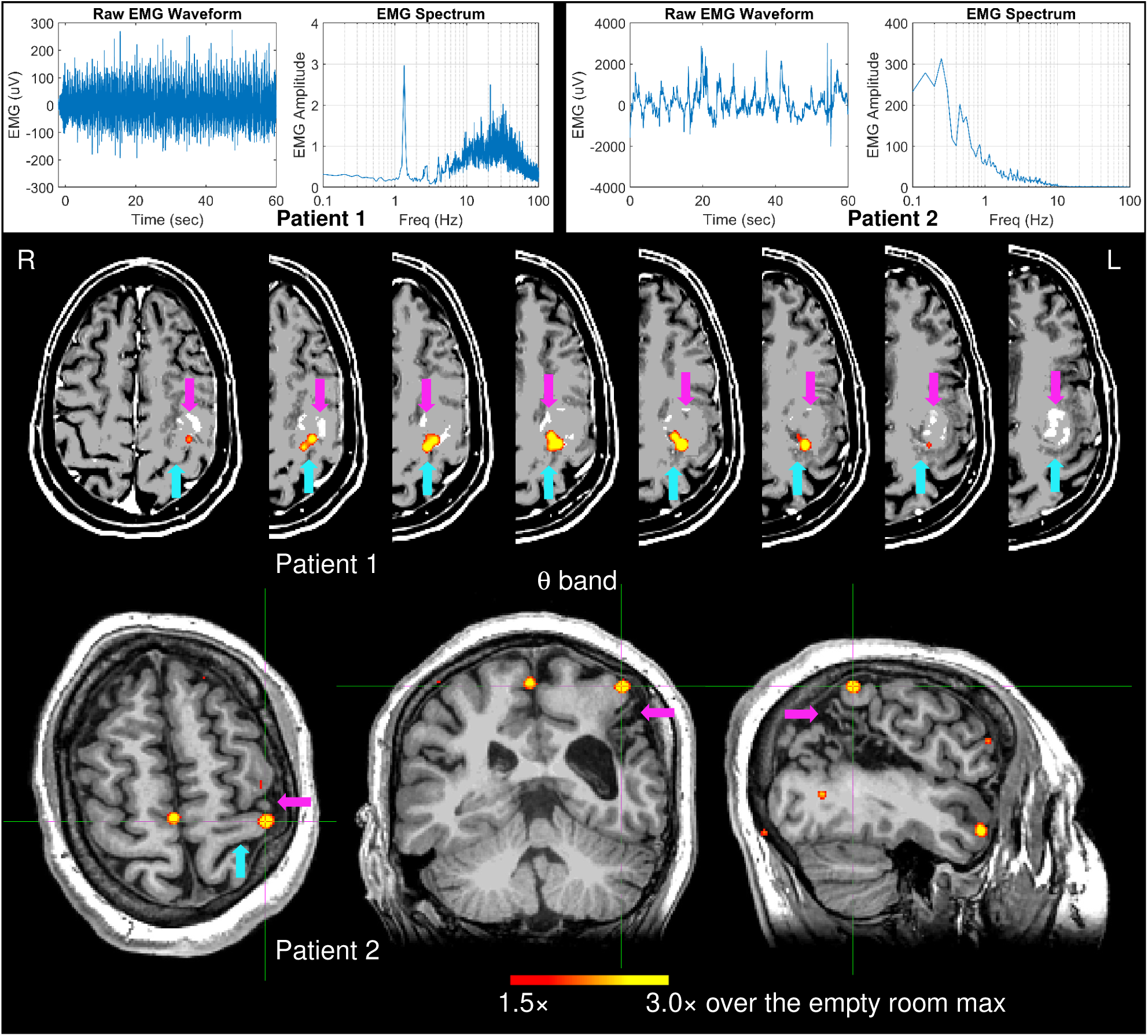
Top panel: EMG waveform and spectrum from two patients with brain lesions when they performed right self-paced (∼1 min) index-finger movements. Bottom panel: Movement-related theta-band EMG-projected MEG source images for the two patients. Primary motor cortex sources contralateral to right self-paced index-finger movements were significant in both patients. In Patient 2, the M1 was also indicated by the crosshairs. The magenta arrows indicate the brain lesions, and the cyan arrows indicate the central sulcus.

Using this presurgical mapping of motor cortex, the neurosurgeon performed a successful gross total resection of the left frontal tumor, avoiding injury to the motor strip. Patient was discharged home the day after surgery, with no neurological deficits.

Patient 2 had chronic cystic encephalomalacia from perinatal stroke, involving the anterior-inferior parietal lobes, with the left much worse than right, primarily involving the lateral left peri-rolandic region and the adjacent more posterior left parietal lobe. Unlike all healthy subjects, Patient 2’s EMG waveform and its spectrum during right index finger self-pace movements (∼ 1 min) (**Fig. 5**, top panel, right plots) show *highly irregular movement patterns and no movement-related frequencies* (fundamental or harmonics). Nevertheless, theta-band EMG-projected MEG source imaging (**Fig. 5** (bottom panel) accurately localized his contralateral left hemisphere M1 source activity just anterior and superior to the cystic encephalomalacia. Note that this patient previously had functional MRI performed on a 3 Tesla MRI at an outside institution, in order to map right hand sensorimotor function, but unlike the MEG study, the fMRI study was unable to localize function. Their fMRI report: *“Right hand motor task: No definite activation of the left sensorimotor area; of note, patient has right hand weakness and right-hand clenching could be performed only for a short duration.”*

Conversely, in the unaffected right M1 cortex of Patient 2, left self-paced finger movement-related frequencies were comparable to healthy subjects and M1 source activity was accurately localized (**Supplementary Fig. S2**, middle and bottom panels). For both the lesioned and non-lesioned hemispheres, theta band M1 source activity was also accurately localized for the 30 sec, but not the 5 sec window (**Supplementary Fig. 2**, top and bottom panels). Owing to the excellent M1 cortex localization in the delta band of healthy subjects, the same analysis was performed in patients for 1-min EMG recordings of finger movements. In both patients, delta M1 activity was significant in both the affected and unaffected hemispheres (**Supplementary Fig. 2**, top and bottom panels).

## Discussion

We developed a novel EMG-projected MEG source imaging technique to localize contralateral M1 cortex activity during repetitive self-paced index finger movements that lasted about 1 minute or less. High-resolution MEG source images were obtained by projecting the MEG brain activity towards the skin EMG signal without trial averaging. In healthy subjects, the EMG-projected MEG approach accurately localized the contralateral M1 regions with good to high efficiency in delta (100.0%), theta (100.0%), and beta (76.9%) bands, whereas efficiency was poor for the alpha (34.6%) and gamma (0.0%) bands. Source imaging of the theta band was even able to localize M1 cortex activity from EMG activity for the first 5 and 30 seconds of self-paced finger movements with good (73.1%) to excellent accuracy (96.2%), respectively. Similarly, in two patients with brain lesions affecting sensorimotor functioning, contralateral M1 cortex theta-band activity was accurately obtained in both cases during 1 min and 30 sec of repetitive finger movements, despite one patient’s highly irregular EMG movement pattern and the absence of movement-related frequencies (fundamental or harmonics). Altogether, these findings extend existing knowledge about M1-muscular couplings with MEG-recorded M1 signals, particularly in low frequency bands, and have translational implications for presurgical localization in patients with brain lesions.

### *Theta*-band MR-CMC

A key finding was that EMG-projected MEG source imaging in the theta band (4-7 Hz) was highly accurate in localizing M1 activity in all cases (100%) of healthy adults and patients alike, which suggests strong movement-related M1-muscle coupling in this band. Movement-related theta band M1-muscle coupling was clearly above the ∼1 Hz movement frequency and its harmonics, indicating that MR-CMC in this band was principally driven by intrinsic motor control. This conclusion aligns with our finding that even when movement patterns were highly irregular without obvious rhythmicity (Patient 2), theta-band M1-muscle coupling still provided accurate localization of M1 cortex activity. Although previous CKC studies reported theta-band SM1 couplings with kinematic features of movements in healthy adults (Jerbi et al., 2007; Piitulainen et al., 2013), movement frequency and its harmonics were primarily driven by movement rhythmicity rather than coherence with muscle activity (Bourguignon et al., 2019). Our results directly show, for the first time, that theta M1-muscle coupling occurs independently of movement rhythmicity. Moreover, our case studies in two patients suggested that EMG-projected MEG source imaging in the theta band during EMG recordings of repetitive finger movements lasting as little as 30 sec is a promising methodology for presurgical M1 mapping even in patients with weakness, numbness, and irregular upper extremity movements.

### *Delta*-band MR-CMC

EMG-projected MEG source imaging also was highly accurate in the delta band (1-4 Hz) in localizing contralateral M1 activity in all healthy subject cases, indicating strong M1-muscle coupling in this band. Unlike the theta band, delta-band activity during our finger movement task includes the fundamental movement frequency (∼1 Hz) and its harmonics. Correspondingly, movement-related CKC in the delta band produces strong SM1 couplings with kinematic features (e.g., speed, velocity, acceleration, and force) at the movement frequency and its harmonics (Bourguignon et al., 2019; Jerbi et al., 2007; Piitulainen et al., 2013). Hence, neuronal sources from CKC appear to be driven by movement rhythmicity, even during repetitive passive movements (Piitulainen et al., 2013). Taken together, neuronal sources subserving delta band M1-muscle couplings appear to support processing of proprioceptive feedback rather than intrinsic motor control (Bourguignon et al., 2019). However, this hypothesis is at odds with a report that delta activity in animals was driven by sensory and strong oscillatory patterns of brain-muscle coupling within the delta band (1.5 – 2.8 Hz) during non-periodic movement behavior (i.e., reaching) (Churchland et al., 2012). Indeed, despite the highly irregular and non-rhythmic self-paced repetitive finger movements of Patient 2, delta-band M1-muscle coupling accurately localized M1 cortex activity. Taken together, these results suggest that delta-band M1-muscle coupling may also be involved in intrinsic motor control processes.

### *Beta*-band MR-CMC

Beta band M1cortex-muscle communication was also found, but contralateral M1 cortex activity reached threshold in only 76.9% of the healthy subject cases. This result is consistent with electrocorticography (ECoG) recordings in epilepsy patients wherein significant coherence was observed between M1 cortex and EMG within the beta band (15–30 Hz) during phasic wrist extensions and flexions (Marsden et al., 2000). Correspondingly, SMC-CMC studies also report beta band (∼20 Hz) M1-muscle coherence using EMG-MEG (Grosse et al., 2002; Pohja et al., 2005), EMG-EEG (Bayraktaroglu et al., 2011; Ushiyama et al., 2017; Yao et al., 2007; Zheng et al., 2016), and EMG-ECoG (Marsden et al., 2000). Yet SMC-CMC is thought to have different neural bases than CKC. In particular, SMC-CMC at ∼20 Hz is a form of SM1-muscular coupling that is linked to the ∼20-Hz component of the sensorimotor mu rhythm (Bourguignon et al., 2019). The mu rhythm is maximal at contraction but is suppressed during movement, which suggests that CMC may not directly involve motor control processes, but rather maintains the current motor state (Engel and Fries, 2010). However, it remains debatable as to whether beta-band MR-CMC is subserved by similar neural process as SMC-CMC, or if the movement-related ∼20 Hz M1-muscle coupling reflects another form of proprioceptive processing like that of CKC (Bourguignon et al., 2019)).

### Weak *alpha*-band MR-CMC

EMG-projected MEG source imaging in the alpha band (∼10 Hz) showed poor localization of M1 activity (34.6%) in healthy subjects. This result aligns with the weak coherence between ECoG recordings at 7-12 Hz and EMG recordings during phasic wrist extension and flexion in epilepsy patients (Marsden et al., 2000). Our finding is also compatible with low M1-muscular coupling in the alpha band in SMC-CMC studies (Bourguignon et al., 2019, 2017; Piitulainen et al., 2015). Weak M1-muscle coupling at ∼10 Hz has been attributed to several factors including event-related desynchronization of the mu alpha rhythm (∼10-Hz) before movement execution (Démas et al., 2020; Pfurtscheller and Lopes da Silva, 1999) and a specific blocking mechanism that prevents the motor pool from synchronizing with descending inputs (Baker et al., 2003).

### Negative *gamma*-band MR-CMC

Gamma-band EMG-projected MEG source imaging failed to localize M1 activity during right and left index finger movement in any of the 13 healthy subjects. This negative result is compatible with our MEG-based brain-computer interface study of decoding hand gestures, for which the gamma-band activity did not contribute to hand-gesture classification accuracy (Bu et al., 2023). In other studies, MEG-based gamma band M1 activity was mainly found post-movement-onset during repetitive finger tapping movements using a seed virtual sensor placed at M1 pre-located from evoked-related finger movement components (Cheyne et al., 2008; Huo et al., 2010). By comparison, the -100 to 0 ms pre-movement-onset window in our study allowed us to examine the early movement execution phase, thereby eliminating post-movement gamma activity for which intrinsic motor control is confounded by other processes (e.g., proprioceptive processing of feedback, maintenance of motor state). Nonetheless, our negative findings should be interpreted with caution, since significant coherence between ECoG and EMG in low (31–60 Hz) and high gamma (61–100 Hz) bands originates from M1 cortex (Marsden et al., 2000). The absence of gamma-band EMG-M1 coupling in our study may be due to lower SNR in the non-invasive MEG gamma band than for invasive ECoG recordings (Bu et al., 2023).

### EMG-projected MEG source imaging approach

Our current results further suggest that the EMG-projected MEG approach requires ∼ 30 sec to 1 min EMG recording time for an accurate localization of M1 in the theta band, which is substantially faster than the conventional movement protocols that last ∼10-20 mins. These short recording times are possible because EMG-projection behaves like a temporal filter to the MEG signals by directly enhancing brain-muscle coupling and substantially eliminating non-muscle related ongoing brain signals. This approach also does not require the movement to be periodic, as EMG-projected MEG was still able to accurately localize M1 in a patient with highly irregular movement patterns and no movement-related rhythms. These findings have important translational applications for accurate M1 cortex localization in presurgical patients who have significant difficulties performing repetitive finger movements because their movements can be slow, weak, and not time-locked to movement-related M1 brain activity. However, more case studies of presurgical patients with sensorimotor dysfunction are needed to fully evaluate our approach.

### Summary

We developed a novel EMG-projected MEG source imaging technique for localizing M1 cortex during EMG recordings of ∼1 minute or less during self-paced repetitive finger movements. The approach was highly accurate in localizing M1 regions in delta, theta, and beta bands, but not in the alpha and gamma bands. Two clinical case studies demonstrated the feasibility and efficacy of this approach in presurgical patients with brain lesions and sensorimotor dysfunction, even in the face of highly irregular and non-rhythmic movement patterns. Our novel EMG-projected MEG source imaging approach in both healthy subjects and presurgical patients also provides insightful information about movement-related brain-muscle coupling above the movement frequency and its harmonics.

## Data Availability

All data produced in the present study are available upon reasonable request to the authors

## Author’s contribution

**M.X. Huang**: Conceptualization, Methodology, Software Development of EMG-projected MEG Source Imaging, Formal Analysis, Supervision on Data Acquisition, Writing-Original Draft, Writing-Review & Editing, Visualization, Project Administration, and Funding Acquisition; **D.L. Harrington**: Conceptualization, Interpretation of Results, Writing-Original Draft, Writing-review and editing, Supervision, Funding acquisition; **A. Angeles-Quinto, Z. Ji**, **A. Robb-Swan, C.W. Huang, Q. Shen, H. Hanson, J. Baumgartner, J. Hernandez-Lucas, T. Song**: Data Curation, Resources, Editing; **S. Nichols, J. Jacobus, I. Lerman, D.G. Baker, R. Rao, M. Bazhenove, G.P. Krishnan**: Supervision, Funding acquisition, Writing-Review & Editing; **R.R. Lee**: Conceptualization, Supervision, Funding acquisition, Writing-Review & Editing.

## Data availability

The data and code that support the findings of this study are available on request from the corresponding author. The data are not publicly available due to privacy or ethical restrictions imposed by School of Medicine, University of California, San Diego.

## Supplementary Materials

**Figure S1:**
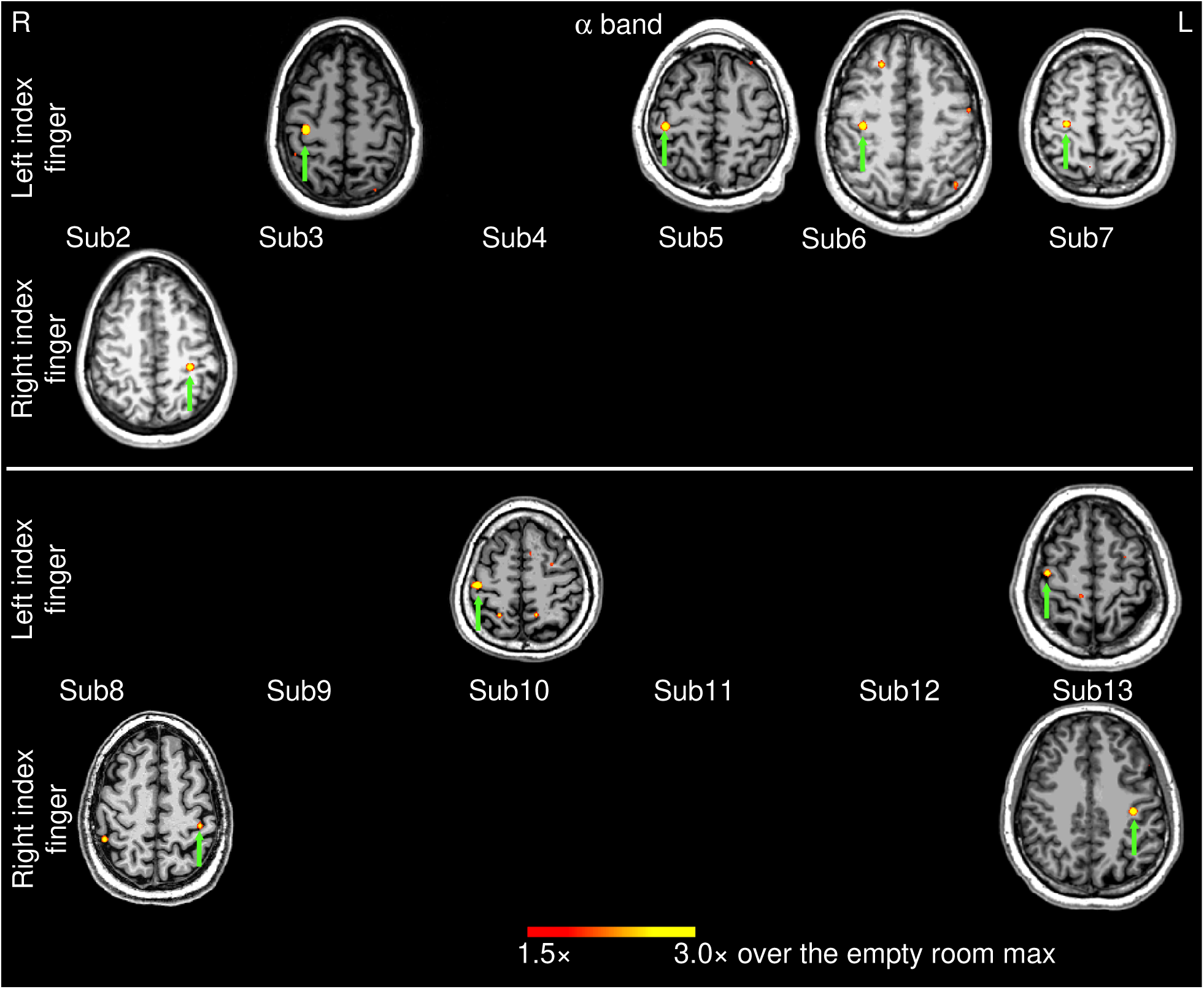
Movement-related alpha-band EMG-projected MEG source images for the remaining 12 healthy subjects. Significant primary motor cortex sources (green arrows) contralateral to left or right self-paced (∼1 min) index finger movements. The color bar shows the activity threshold at 1.5× of the empty room maximum value, and saturation level at 3.0×. In 6 cases, contralateral primary motor source activity at or above the threshold was not observed. Sub = subject

**Figure S2:**
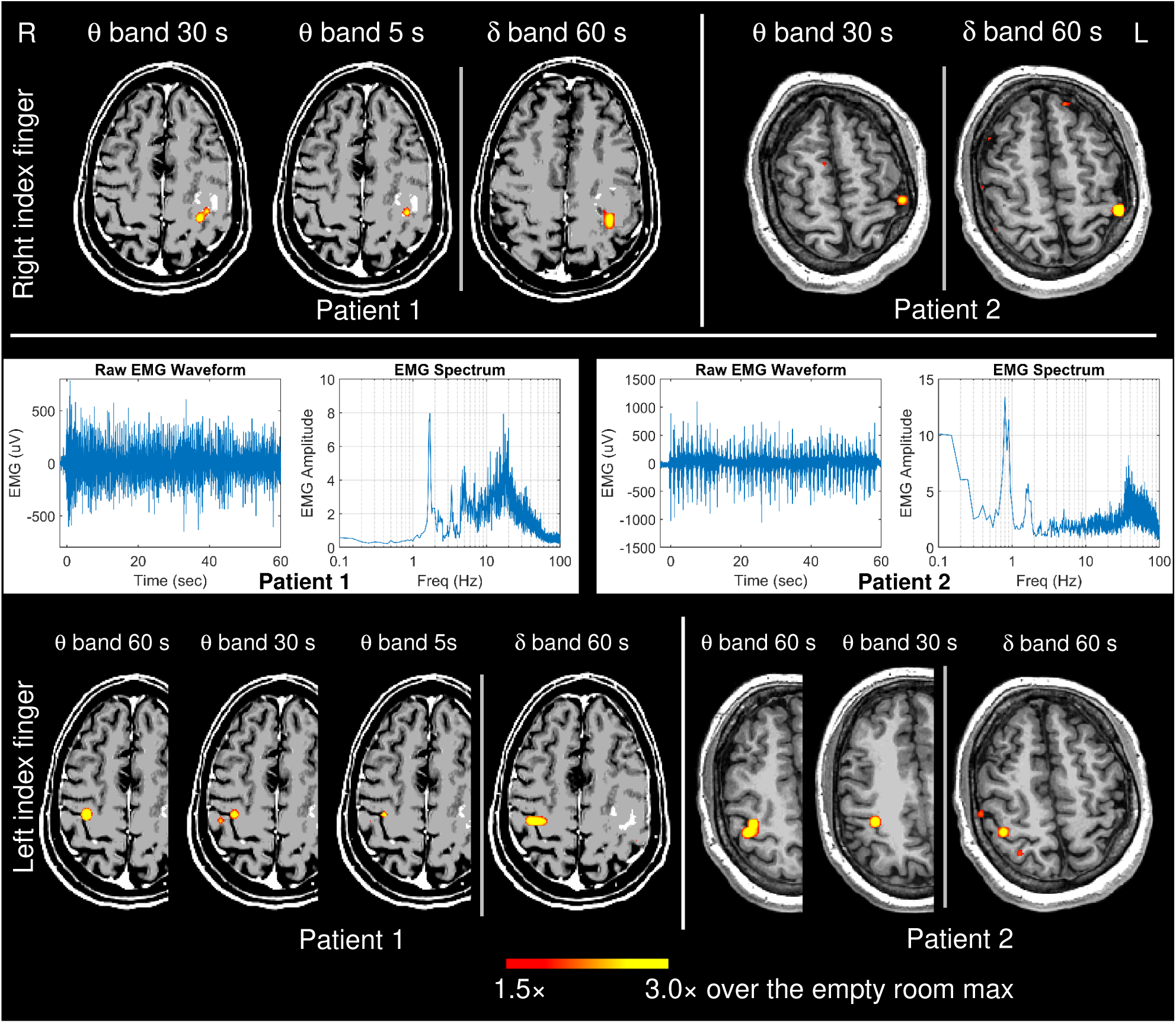
This figure provides supplementary results to Figure 5 in the main text. Top Panel: EMG-projected MEG source images of M1cortex in the affected left hemispheres due to right finger movement (affected hand) from the two clinical patients in theta and delta bands. In the theta band, M1 was localized in 30 sec and 5 sec time windows in Patient 1, but only the 30 sec time window in Patient 2. Delta-band M1 cortical activity (1 min EEG recordings) in the damaged hemisphere was localized in both patients. Middle Panel: the EMG waveforms and spectra from the left index finger movement (unaffected hand). Bottom Panel: EMG-projected MEG source images of M1 in the unaffected right hemispheres due to left finger movement from the patients in theta and delta bands. In Patient 1, theta-band M1 cortex activity was localized in the 60 sec, 30 sec and 5 sec time windows. In Patient 2, theta-band M1 activity was localized only in the 60 sec and 30 sec time windows. Delta-band M1 activity (1 min EEG recordings) was localized in the unaffected hemisphere in both patients.

